# Virtual brain twins for stimulation in epilepsy

**DOI:** 10.1101/2024.07.25.24310396

**Authors:** Huifang E. Wang, Borana Dollomaja, Paul Triebkorn, Gian Marco Duma, Adam Williamson, Julia Makhalova, Jean-Didier Lemarecha, Fabrice Bartolomei, Viktor Jirsa

**Affiliations:** Aix Marseille Univ, INSERM, INS, Inst Neurosci Syst, Marseille, 13005, France; APHM, Epileptology and Clinical Neurophysiology, Department,Timone Hospital; Marseille, 13005, France; Aix Marseille Univ, CNRS, CRMBM; Marseille, 13005, France; APHM, Timone University Hospital, CEMEREM; Marseille, 13005, France; Epilepsy Unit, IRCCS E. Medea Scientific Institute, Via Costa Alta 37, Conegliano, Treviso 31015, Italy

**Keywords:** Personalized medicine, Brain modelling, Epilepsy, Stimulation, Stereo-EEG, Temporal interference, Diagnosis and treatment

## Abstract

Estimating the epileptogenic zone network (EZN) is an important part of the diagnosis of drug-resistant focal epilepsy and plays a pivotal role in treatment and intervention. Virtual brain twins based on personalized whole brain modeling provides a formal method for personalized diagnosis by integrating patient-specific brain topography with structural connectivity from anatomical neuroimaging such as MRI and dynamic activity from functional recordings such as EEG and stereo-EEG (SEEG). Seizures demonstrate rich spatial and temporal features in functional recordings, which can be exploited to estimate the EZN. Stimulation-induced seizures can provide important and complementary information. In our modeling process, we consider invasive SEEG stimulation as the most practical current approach, and temporal interference (TI) stimulation as a potential future approach for non-invasive diagnosis and treatment. This paper offers a virtual brain twin framework for EZN diagnosis based on stimulation-induced seizures. This framework estimates the EZN and validated the results on synthetic data with ground-truth. It provides an important methodological and conceptual basis for a series of ongoing scientific studies and clinical usage, which are specified in this paper. This framework also provides the necessary step to go from invasive to non-invasive diagnosis and treatment of drug-resistant focal epilepsy.

## 1 Introduction

To obtain an accurate diagnosis in the most complex cases of drug-resistant focal epilepsy, the clinical routine includes invasive stereo-EEG (SEEG) implantation to estimate the epileptogenic zone network (EZN), which are key elements for a successful treatment [1, 2]. SEEG has become one of the principal techniques for delineating EZNs [3, 4]. In the past 15 years, several data analysis methods for quantifying EZNs were proposed based on the spectral analysis of SEEG signals [5, 6]. Beyond pure data-driven analysis approaches, several methods linking mechanistic models and data analysis have been developed [7–12], formally exploiting causal hypotheses within an inference framework. We developed a workflow for the estimation of a patient’s EZN using personalized whole brain models, called the virtual epileptic patient (VEP), [13–16]. The VEP workflow was evaluated retrospectively using 53 patients with 187 spontaneous seizures and is now being evaluated in an ongoing clinical trial (EPINOV) with an expected 356 prospective patients with epilepsy [14, 15]. The virtual brain twin concept has been proposed based on the VEP workflow and was extended to various brain disorders [17]. Virtual brain twins are personalized, generative and adaptive brain models based on data from an individual’s brain for scientific and clinical use. In this study, we introduce the first high-resolution virtual brain twin workflow, specifically designed for estimating the EZN using a stimulation paradigm.

During SEEG, direct electrical stimulation can be used to map brain function as well as to provoke seizures for diagnosis. SEEG stimulation-induced seizures (at 1 Hz or 50 Hz, usually with pulses of 1 ms at 1-3 mA) are an important tool for localizing the EZ and are also associated with a better post-surgical outcome [18–20]. First, we propose a personalized whole brain model, i.e a virtual brain twin, dedicated to assessing stimulations performed through SEEG electrodes. We build a high-resolution brain model for an individual patient and compute the electric fields induced by SEEG stimulation. We simulate the brain responses induced by SEEG stimulation and estimate the EZN based on the induced seizures. As the ground truth is known for the simulated data, we can formally validate the EZN estimation. Then, we evaluate the capacity of our approach to translate from invasive stimulation and recording via SEEG to non-invasive procedures using scalp-EEG recordings and transcranial electrical stimulation techniques, notably temporal interference (TI). The recently developed TI has the capacity of reaching deeper structures than conventional transcranial direct/alternating current stimulation (tDCS/tACS) [21, 22]. Thus combining the advantages of tDCS and DBS [23], TI stimulation [24] is both non-invasive and focal and can stimulate deep structures of the brain. TI exploits the insensitivity of brain tissue to electric stimulation at high frequencies (above 500 Hz) [24]. When two slightly detuned high frequency electric fields are applied at different locations, their interference provides an envelope modulation at a lower frequency, equal to the difference between the two electric field frequencies Δ*f* (normally below 150 Hz), and acts as an effective stimulation at a desired targeted location. Thus the obtained low frequency modulation is relevant to functional dynamics of the brain.

In this study, we provide a workflow estimating the EZN based on personalized high-resolution virtual brain twin under a stimulation paradigm. We develop a pipeline that can 1) build a personalized high-resolution brain model for either SEEG or TI stimulation; 2) estimate the EZN from stimulation-induced seizures, and validate it by simulated data; and 3) refine the decision-making process by integrating multiple recording modalities, such as scalp-EEG and SEEG. This research provides a necessary step for 1) a series of scientific and clinical studies, such as optimization of stimulation parameters for diagnosis and treatment; 2) moving from invasive to non-invasive diagnosis and treatment of drug-resistant focal epilepsy; 3) natural integration of multiple functional data modules.

## 2 Results

### 2.1 VEP-stimulation workflow

We built a high-resolution virtual brain twin workflow to estimate the EZN using stimulation techniques. An EZN is defined as the tissue responsible for generating seizures and may involve distant brain areas characterized by altered excitability [1]. Our model-based high-resolution workflow using perturbation techniques for diagnostic EZ mapping is shown in Fig. 1 and the detailed flowchart is shown in Fig. **??**. First, high-resolution full brain network models (in the middle panel of Fig. 1) were established using patient-specific data from patients with epilepsy. The structure of the model was defined by the detailed surface of the cortex and the subcortical volumes. The heterogeneous structural connectivity between brain regions through white matter fibers was estimated from diffusion-weighted MRI. The cortical surface data was generated from T1-weighted MRI using the Freesurfer software package, resulting in surfaces with 20284 vertices with a vertex area of about 10 *mm*^2^. We simulated the time series on the patient’s specific structural scaffold using the phenomenological Epileptor model [25], a system of differential equations that can describe seizure initiation, propagation, and termination, resulting in electrophysiological seizure like events. The spatial domain of the Epileptor is given by the high-resolution network of neural masses, thus seizures can propagate locally across neighboring vertices of the cortex and globally through white matter fiber connections [7]. The personalized modelling parameters can be inferred from the spontaneous seizure recording [15].

**Fig. 1.**
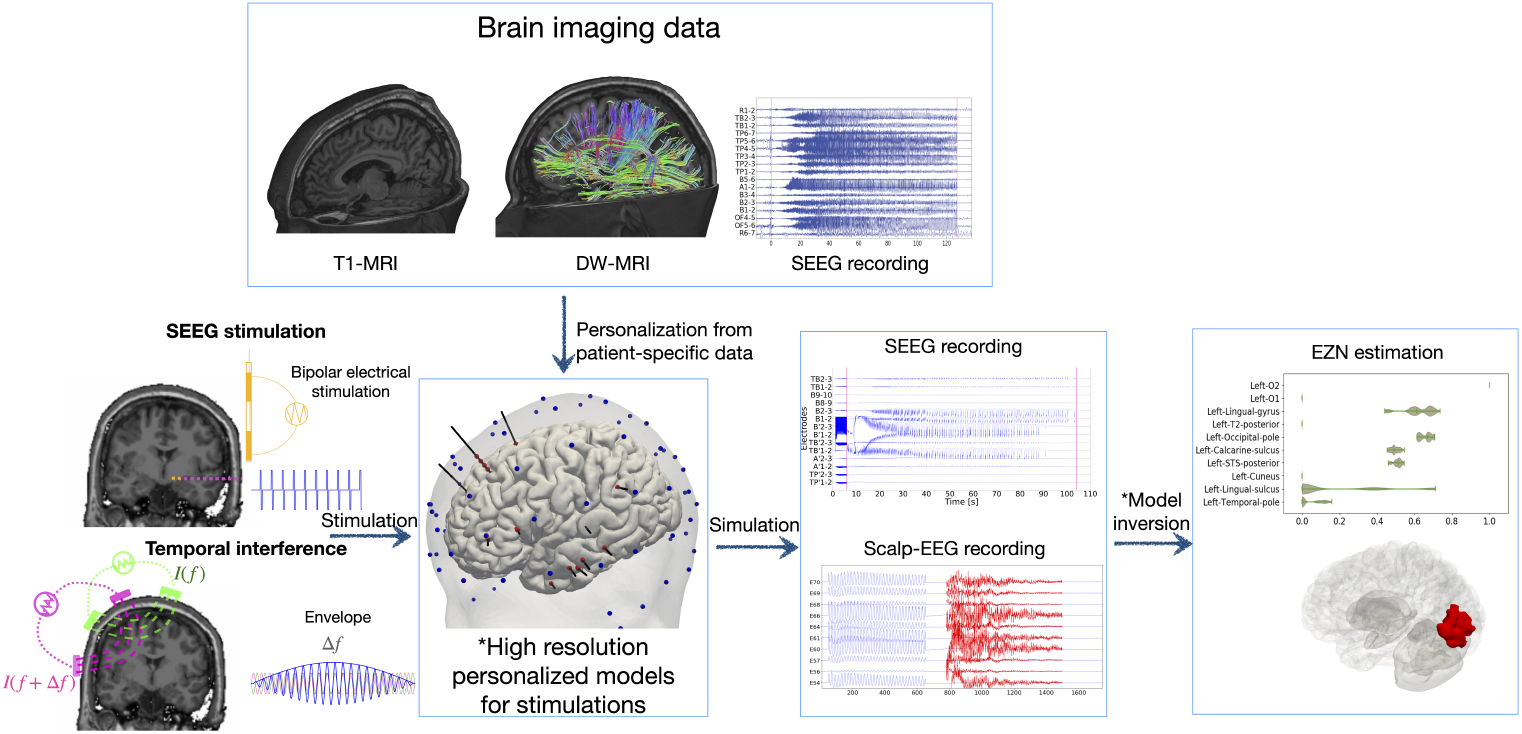
The workflow of the virtual brain twin for estimating the epileptogenic zone network (EZN) using stimulation techniques. A personalized high-resolution model (in the middle) is based on individual brain geometry extracted from T1-weighted MRI and structural connectivity from tractography on diffusion-weighted MRI data (in the top). High resolution virtual brain models simulate neural source activity with spatial resolutions of about 10 *mm*^2^. The modelling parameters are inferred from the spontaneous SEEG recordings (in the top). We illustrated two types of stimulation: SEEG and Temporal interference (TI), to induce seizure activity. SEEG stimulation (in the left top) uses bipolar stimulation in which two electrodes are used: one serves as the cathode and the other as the anode. The electric current flows between two electrodes, which is defined by current amplitude, pulse width and frequency. TI stimulation (in the left bottom) applies two current sources simultaneously via electrically isolated pairs of scalp electrodes (green and pink) at kHZ frequencies *f* and *f* + Δ*f*. The currents generate oscillating electric fields which results in an envelope amplitude that is modulated periodically at Δ*f*. The electric field influences the brain activity which can be generated by the high-resolution personalized whole-brain model (in the middle). The red and blue dots represent SEEG and scalp-EEG electrodes, respectively. The simulated source activity can be mapped onto the corresponding SEEG and scalp-EEG signals (the 3rd column in the bottom panel), through the gain matrices, which are constructed based on the locations of SEEG/scalp-EEG electrodes relative to the source vertices. The red curves on the scalp-EEG recordings are plotted using a different scale to visualize the signals following the high-amplitude signals induced by TI stimulation. By utilizing data features extracted from SEEG and scalp-EEG signals, Bayesian inference methods can estimate a posterior distribution of epileptogenic values, suggesting the potential EZN (in the right).

In this study, we developed the first high-resolution virtual brain twin designed for stimulation. First, we calculated the electric fields induced by both stimulation methods (in the left panel of Fig. 1). For the SEEG stimulation, two SEEG electrodes serve as cathode and anode to generate bipolar stimulation and electric current flow is defined by current amplitude, pulse width and frequency (illustrated in top left of Fig. 1). The perturbation effect was applied to the vertices of the high-resolution surface through the SEEG-to-source mapping, i.e. a higher perturbation effect was mapped on the vertex that was at shorter distance from the stimulation electrodes. For the TI stimulation, we calculated the envelope amplitude of the TI electric field for each vertex whose envelope is modulated at the frequency Δ*f* (illustrated in bottom left of Fig. 1). Both types of stimulation generated an accumulative increase in the state variable *m* of the Epileptor-Stimulation model (eq. 1). The slowly evolving state variable captures changing tissue properties under electric stimulation. When *m* reaches a given threshold, the model undergoes a transition into the seizure state. A post-SEEG implantation CT scan is used to localize SEEG contacts and co-register them with the structural scaffold. Scalp-EEG electrodes were placed on the scalp using the standard international 10-5 system to place scalp-EEG electrodes. This high-resolution model allows us to consider detailed electric dipoles, generated by neural activity, for building high-fidelity forward solutions. The gain matrix maps the activity from the neural sources (located at vertices of the surface and subcortex) to the electrodes (SEEG/scalp-EEG), based on their orientation and distance.

Model inversion estimates patient-specific brain model parameters, especially epileptogenicity and global network scaling using Hamiltonian Monte Carlo (HMC) sampling techniques from Bayesian inference methods [10, 15]. The estimation is based on the structural brain scaffold, modelled seizure dynamics, and the data feature extracted from scalp-EEG and/or SEEG seizure recordings (the 3rd column in the bottom panel in Fig. 1). The model inversion used a noninformative prior in which all the brain regions followed the same prior distribution (the prior assumption is all regions are healthy). The result is the posterior probability from which the EZN is identified. We also introduced multimodal inference for simultaneous SEEG/scalp-EEG to infer the EZN.

### 2.2 Virtual brain twins for SEEG stimulation

We used the data from a right-handed female patient in her 20s diagnosed with left occipital lobe epilepsy to illustrate how to use our workflow. We first extracted the brain geometry, the structural connectivity matrix and the source-to-sensor mapping from the T1-MRI, diffusion-weighted MRI, and post-SEEG-implantation CT scans for this patient. We built the whole brain neural mass model based on this anatomical information. Then we ran the HMC algorithms for three spontaneous seizures recorded from this patient (one of the three seizures is shown in Fig. 2 (**A**)). We pooled the posterior distribution from the three seizures, in which the left lateral occipital cortex (region O2 (in VEP atlas [26]) was consistently identified as part of the EZ. Then we projected the left O2 onto both preoperative and postoperative T1-MRI slides and 3D brain meshes (in Fig. 2(**C-E**)) to compare its extent with the size of the surgical resection. The patient underwent resective surgery, in which a large portion of the left O2 was removed, and was almost seizure free after surgery (surgery outcome class of Engel II [27]). Based on this result, we built the high-resolution whole brain model for SEEG- and TI-stimulation with the hypothesis that the left O2 was the EZ of the patient.

**Fig. 2.**
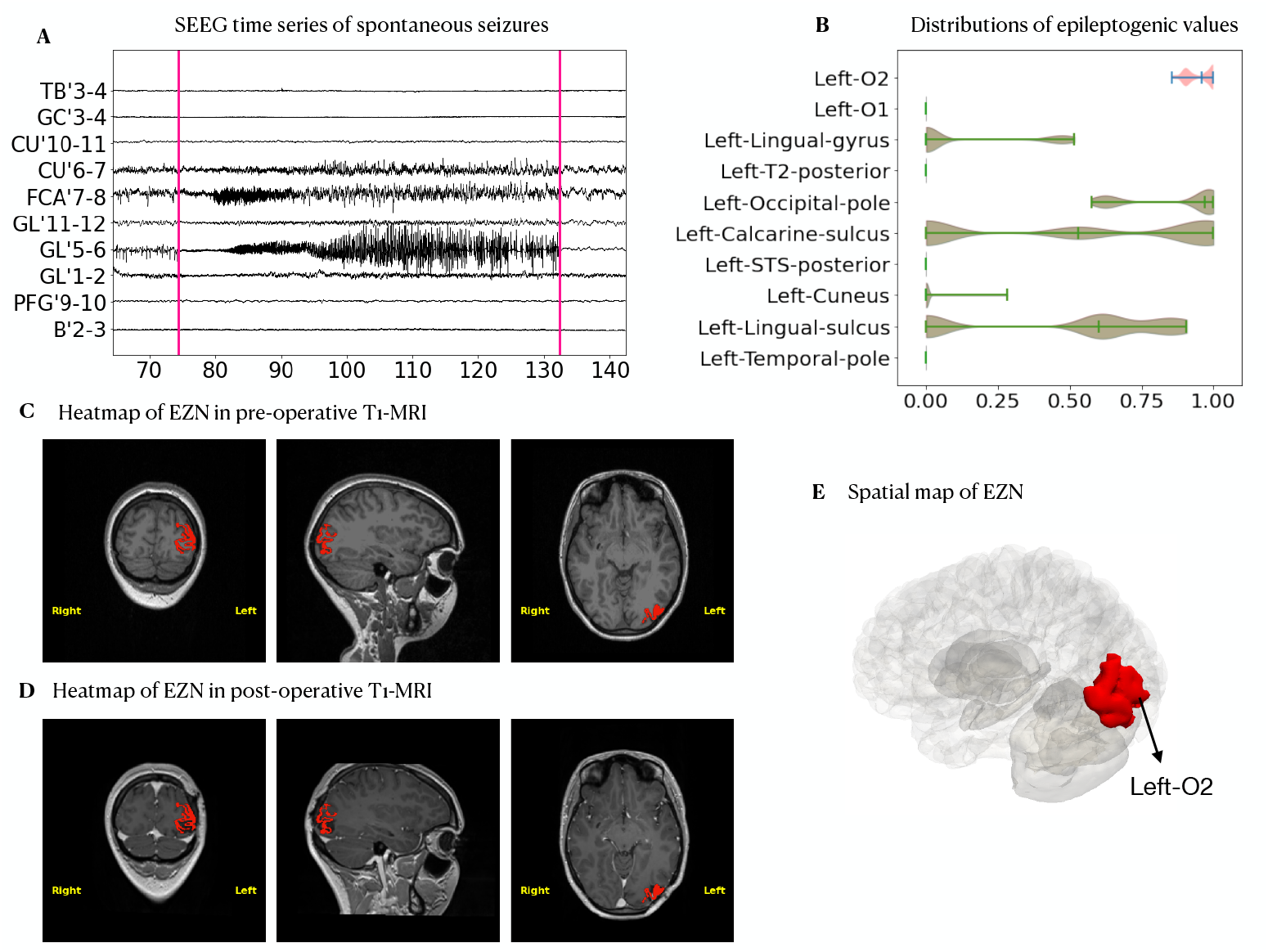
VEP diagnostic mapping for spontaneous seizures. (**A**) Selected SEEG recordings from one seizure in a female patient in her 20s. (**B**) Posterior distribution of the epileptogenic values (EV) (higher value indicates higher probability of seizure) for ten selected regions obtained from the HMC pipeline. (**C**) Heatmap of the left O2 identified by VEP (in red) shown in a preoperative T1-MRI. (**D**) Heatmap of the left O2 identified by VEP (in red) shown in a postoperative T1-MRI. (**E**) The left O2 (in red) was projected on the patient’s 3D meshes.

To model direct electrical stimulation by SEEG electrodes in the brain, we first mapped the contribution of SEEG stimulation current to brain source activity. We retrieved the contribution from a pair of electrodes, in which the perturbation is applied to the brain regions based on the sensor-to-source mapping matrix [15]. Then we calculated the effect of the bipolar pulse stimulation (50 Hz for a duration of 3.5 s on bipolar GL’5-6 electrode leads) on each vertex of the cortex mesh (Fig. 3**A**). We used the Epileptor-Stimulation model on each of the 20,284 vertices. The stimulation of the Epileptor leads to an accumulation in a state variable *m* that pushes the brain to seize (Fig. **??**). The seizures were located around the left O2 regions. From these simulated neural source signals (Fig. 3**B** and Movie S1), we obtained SEEG signals (Fig. 3**C**) using the source-to-sensor gain matrix while considering the orientation of dipoles.

**Fig. 3.**
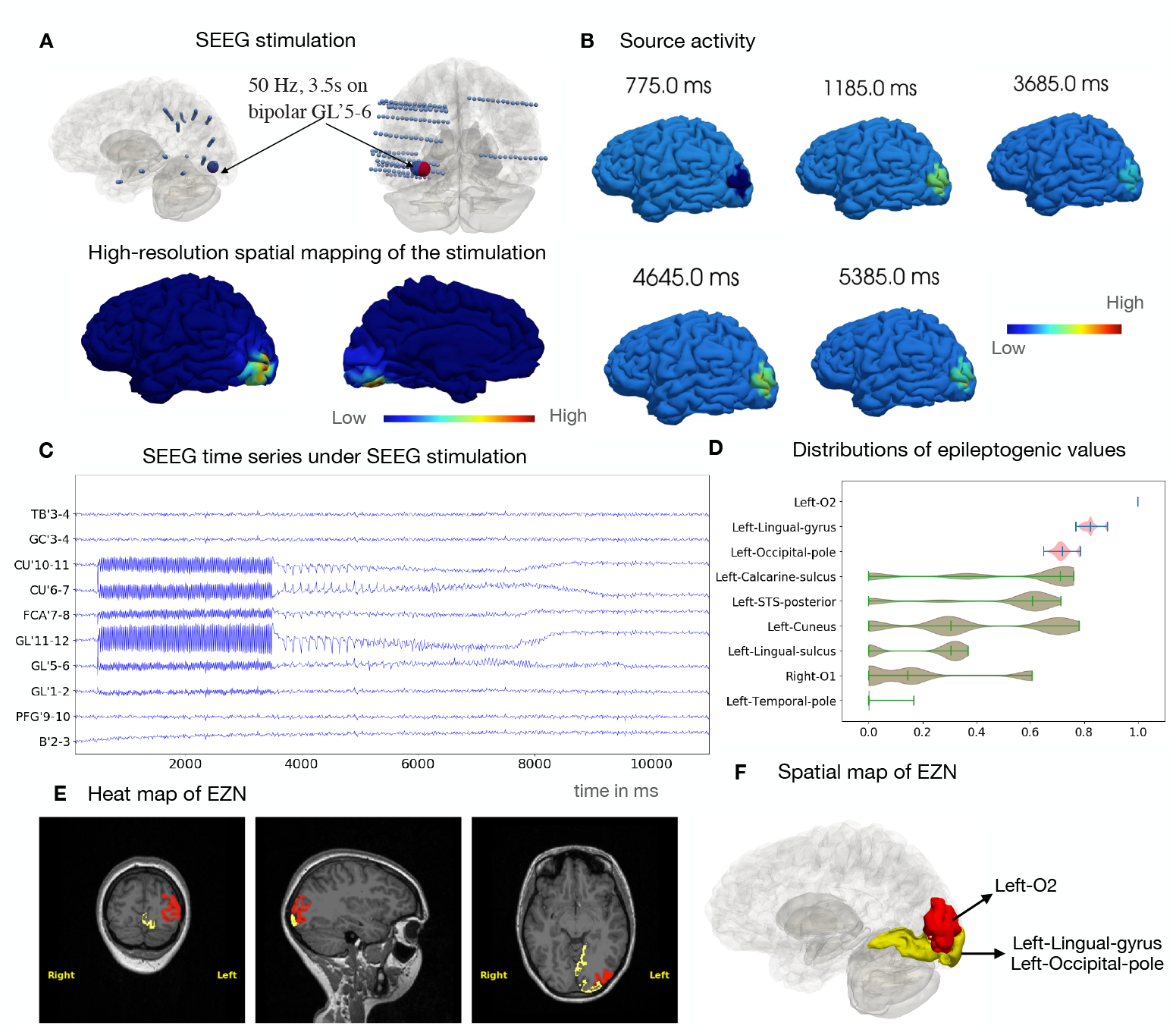
Estimating EZN for SEEG stimulation induced seizure. (**A**) GL’5-6 (large blue/red sphere) is the stimulated contact in the left occipital lobe, using a bipolar pulse stimulation (50 Hz for a duration of 3.5 s with pulse duration 1 ms) (Top). The spatial mapping of the maximum of stimulation values (shown in the colorbar from low (left) to high (right) values) for each vertex of the brain from sensor GL’5-6 (bottom). (**B**) Neural source activity is shown on the cortical mesh at 5 different time points. The seizures are located around the left O2 region of the VEP atlas. (**C**) Selected simulated SEEG time-series from a SEEG stimulation-induced seizure. (**D**) Posterior distribution of the EV (higher value indicates higher chance for seizure) of 9 selected regions obtained from HMC sampling. Red regions indicate highest chance of being the EZ, other areas are in green. The region of the highest EV posterior distribution is the left O2 in red shown in T1-MRI (**E**) and 3D brain in (**F**). The two regions (the left lingual gyrus and the left occipital pole) are shown in yellow in (**E, F**).

These synthetic SEEG recordings (Fig. 3**C**) were used for model inversion, to estimate the EZN. We obtained the left O2 with the highest EVs out of all the samples according to the posterior distribution from the HMC algorithm (Fig. 3**D**). The two neighboring regions (the left-lingual-gyrus and the left-occipital-pole) were the second-group of candidates that could belong to the EZN. The heatmap projected on the patient’s T1-MRI revealed the spatial mapping of the EZN (sagittal, axial and coronal view images shown in Fig. 3**E**). The spatial mapping of these three regions in 3D brain is highlighted in red and yellow in Fig. 3**F**.

### 2.3 Virtual brain twins for TI stimulation

We demonstrate now how the pipeline works on TI stimulation. First, we used high resolution modeling for TI stimulation. We calculated the TI electric field, which is projected onto the cortical surface by stimulating two pairs of scalp-EEG electrodes PPO3-PPO5 and P5h-PO5h [28] in Fig. 4**A**) at the frequencies of 1000 Hz and 1005 Hz, respectively. The stimulation amplitude is determined based on the assumption that the components of the field that have the greatest impact are those that are parallel to the neurons’ axons [29]. We used the amplitude of the TI electric field as input to the Epileptor model on each vertex of the cortical mesh (Fig. **??**). We simulated the brain source signals on each vertex of the whole brain by considering both global and local connectivity (Fig. 4**B** and Movie S2). We then mapped the signals on the vertices of the cortical surface onto the scalp-EEG signals (Fig. 4**C**) using the source-to-sensor mapping matrix.

**Fig. 4.**
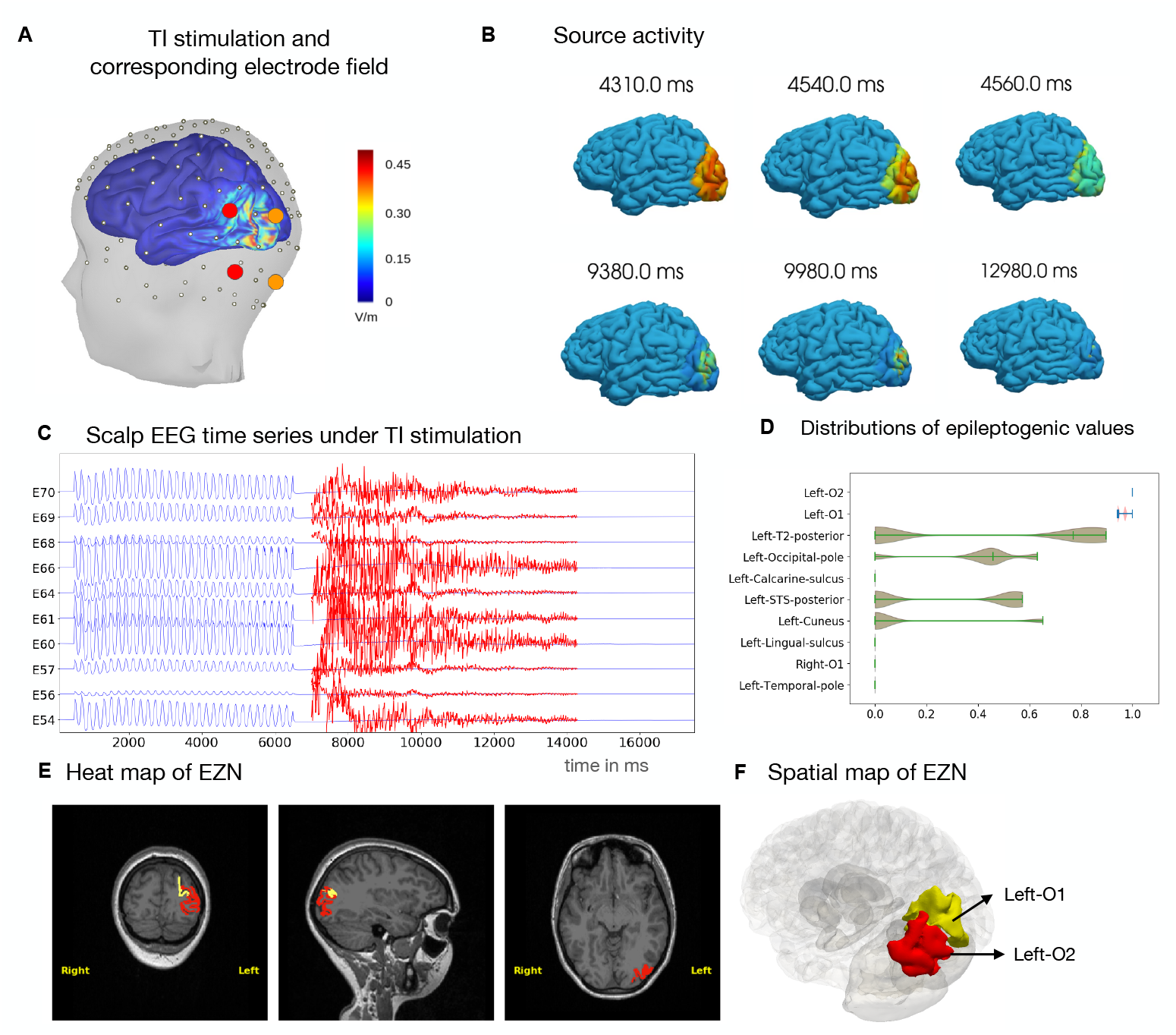
Estimating EZN from TI stimulation induced seizure. (**A**) The electric field of TI stimulation by two pairs of scalp-EEG electrodes (shown in red and orange) based on the 10-5 international reference system, using an extended scalp-EEG cap from SIMNIBS [28]. We applied a frequency of 1000 and 1005 Hz for the first and second electrode pairs, respectively. The spatial distribution of the peak activity of the TI envelope is colored in the 3D brain. (**B**) Seizure dynamics were simulated using the Epileptor-Stimulation model through the TI stimulation. Neural activity is shown on the cortical mesh at 6 different time points. The seizures are located around the left O2 regions of the VEP atlas. (**C**) Selected simulated scalp-EEG time-series from TI stimulation-induced seizure. The scaled-up time series during the seizure period is shown in red. (**D**) Posterior distribution of the EV (higher value indicates higher chance for seizure) for 10 selected regions obtained from the HMC sampling. Red regions indicate highest chance of being the EZN; the others are in green. The region of the highest EV posterior distribution is the left O2 in red shown in T1-MRI (**E**) and 3D brain in (**F**). The second region (Left O1) is shown in yellow in (**E, F**).

We extracted the data features from these synthetic scalp-EEG recordings (Fig. 4**C**) as input to the HMC model inversion to estimate the epileptogenic zone. From the posterior distribution of EVs, we obtained the left O2 as first and the left O1 as the second candidate belonging to the EZN (Fig. 4**D**). The heatmap of these two regions projected on the patient’s T1-MRI shows the spatial mapping of the EZN (sagittal, axial and coronal view images shown in Fig. 4**E**) and the 3D brain in Fig. 4**F**, where the left O2 is in red and the left O1 in yellow.

### 2.4 Multimodal inference

For patients with multiple seizure recordings or simultaneous multiple modality recordings (here we used SEEG and scalp-EEG as examples), we integrate both modalities into the model inversion algorithm to combine the information from multiple functional data sources in the parameter estimation process and potentially improve clinical decision making. Here we demonstrated four possible cases in which the EZN estimation from multimodal functional recordings, such as scalp-EEG and SEEG, were integrated. The first case estimated the EZN from simultaneous scalp-EEG and SEEG recordings under SEEG stimulation (Fig. 5**A** and Fig. **??A**). Here we mapped the same source signals to both the SEEG and scalp-EEG sensors in both the simulation and during model inversion. Two added values come about as a result of doing simultaneous SEEG and scalp-EEG. First, the scalp-EEG provides a whole brain sampling although it mainly measures weak cortical surface signals in contrast to the SEEG, which only samples partial brain space with relatively strong signals from deep structures as well. Second, this pipeline provides a way to evaluate the roles of the scalp-EEG so that we could design a less invasive SEEG implantation in the future. The posterior of the EVs from a simultaneous model inversion showed the left O2 as the EZ (Fig. **??A**). The second case estimated the EZN from simultaneous scalp-EEG and SEEG recordings under TI stimulation (Fig. 5**B** and Figure **??B**). The posterior of the EVs from the multimodal model inversion showed the left O2 as the EZ and the left STS-posterior as a second candidate. In the third case, we utilized the multimodal model inversion module on SEEG recordings from SEEG stimulation (Fig. 5**A**) and scalp-EEG recording from TI stimulation (Fig. 5**B**). The posterior of the EVs from the multimodal model inversion showed the left O2 as the EZ (Fig. **??C**). Notice that, in this case, the functional SEEG and scalp-EEG data here were not simultaneous recordings. The results would be valid only with an assumption that the HMC model inversion sampled all feasible initial conditions. For comparison, in the forth case, we pooled the posterior distributions of the EVs from the SEEG (in Fig. 3**D** under SEEG stimulation) with the EVs from the scalp-EEG (in Fig. 4**D** under TI stimulation). We obtained the left O2 as the intersection of the two distributions to be the only brain region in the EZN (Fig. **??D**).

**Fig. 5.**
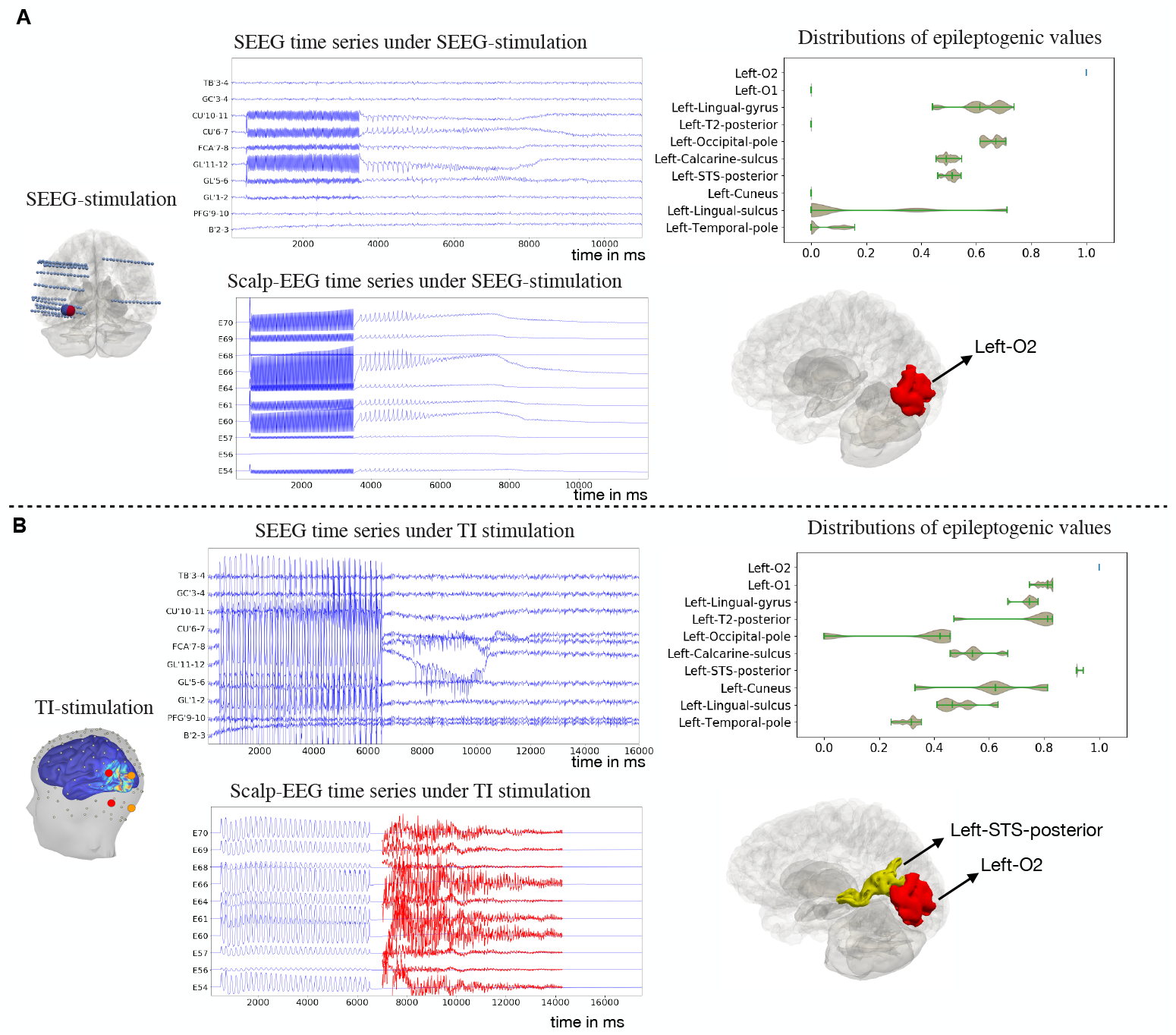
Multimodal inference for EZN estimation from simultaneous SEEG and scalp-EEG recordings. (**A**) We combined ictal recordings from SEEG (middle top) and scalp-EEG (middle bottom) induced by SEEG stimulation (left), using multimodal inference to obtain the distribution of EVs (right top). The results were mapped in 3D. In this case, we obtained the ground-truth left O2 in red. (**B**) We combined ictal recordings from SEEG (middle top) and scalp-EEG (middle bottom) induced by TI stimulation (left), using multimodal inference to obtain the distribution of EVs (right top). The results were mapped in 3D. In this case, we obtained the ground-truth left O2 in red and additional brain region as left STS-posterior in yellow.

### 2.5 Validation of the pipeline

To validate that the pipeline can be easily used in other patients, we added the results of a second patient with left frontal lobe epilepsy. The patient underwent resective surgery resulting in seizure freedom (surgery outcome class of Engel I [27]). Eight brain regions including the orbito frontal, anterior-cingulate, F1 lateral prefrontal, F2 rostral, SFS rostral, frontal pole and F1-mesial-prefrontal cortex in the left hemisphere were resected. We built the high-resolution whole-brain model for two types of perturbations under the hypothesis that the left F1 lateral prefrontal is the EZ. To model direct electrical stimulation by SEEG electrodes in the brain, we calculated the effect of the bipolar pulse perturbation (50 Hz, for a duration of 3 s on bipolar PM’3-4 electrode leads) (Fig. 6**A**). The seizure was induced around the left F1 lateral prefrontal cortex. From the simulated neural source signals (Fig. 6**B** and Movie S3), we obtained simultaneous SEEG and scalp-EEG signals (Fig. 6**C**) using the source-to-sensor gain matrix. These synthetic SEEG and scalp-EEG signals during ictal period served as input for multimodel inference to estimate the EZN. We obtained the left-F1-lateral-prefrontal cortex as the region with the highest EV from the HMC algorithm (Fig. 6**D**,**E**). The two neighboring regions (the left-F1-measial-prefrontal and the left-middle-frontal-sulcus) were identified as secondary candidates that could belong to the EZN (Fig. 6**D**,**E**).

**Fig. 6.**
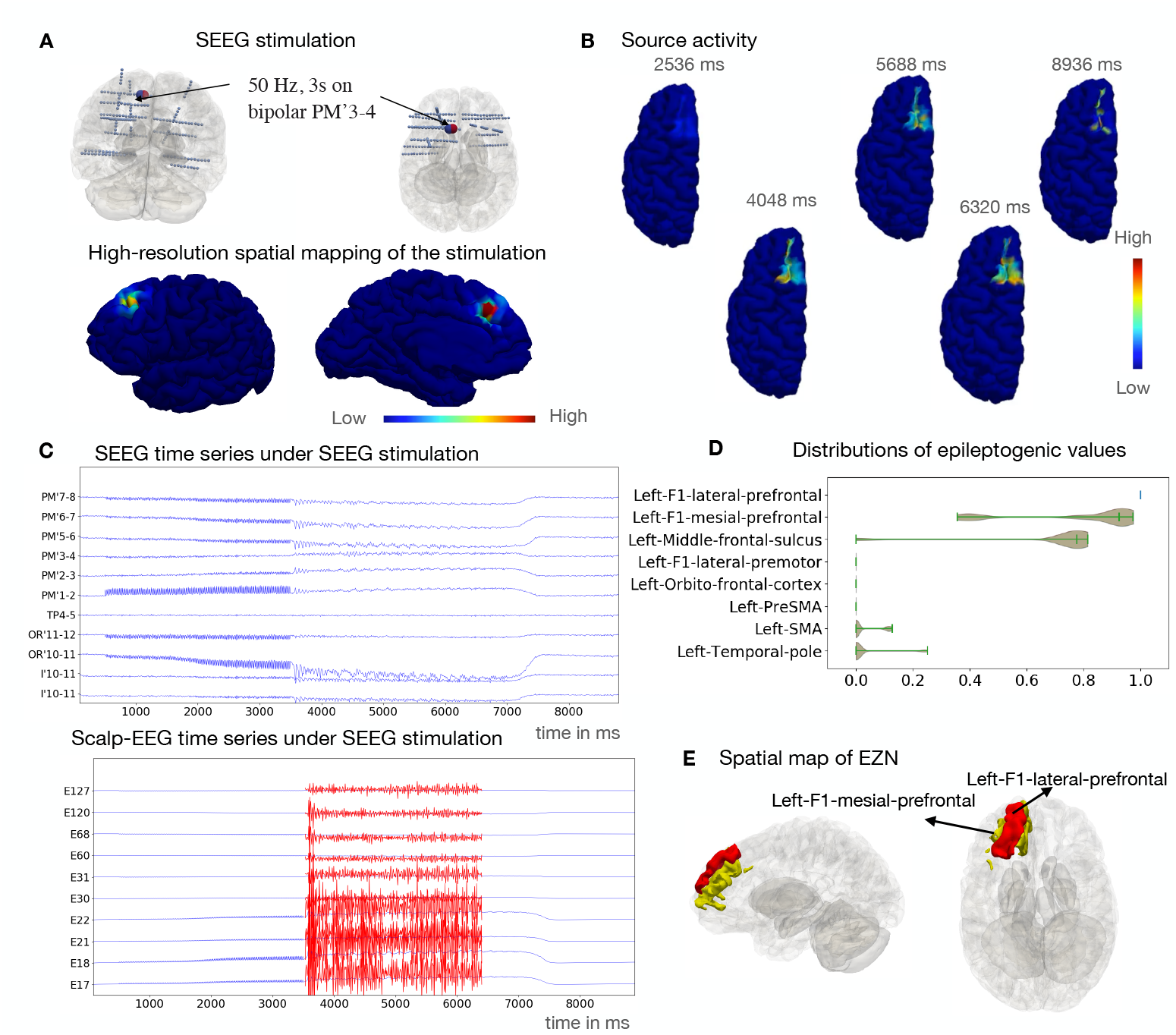
Estimating EZN from SEEG stimulation for a second patient with frontal lobe epilepsy. (**A**) PM’3-4 (large blue/red sphere) is the stimulated contact in the left frontal lobe, using a bipolar pulse stimulation (50 Hz for a duration of 3 s with pulse duration 1ms) (Top). The spatial mapping of the maximum of stimulation values (shown in the colorbar from low (left) to high (right) values) for each vertex of the brain from sensor PM’3-4 (bottom). (**B**) Seizure dynamics were simulated using the Epileptor-Stimulation model and induced through SEEG stimulation. Neural activity is shown on the cortical mesh at 5 different time points. (**C**) Selected simulated SEEG time-series (top) and EEG time-series (bottom) from SEEG stimulation-induced seizure. The scaled-up time series during the seizure period is shown in red. (**D**) Posterior distribution of EVs (higher value indicates higher chance for seizure) for 8 selected regions obtained from the HMC sampling when analyzing simultaneously SEEG and scalp-EEG. (**E**) The highest chance of being the EZN as left-F1-lateral-prefrontal in red mapped in 3D brain and left-F1-mesial-prefrontal cortex and left-middle-frontal-sulcus in yellow.

We also illustrated the results of this frontal lobe epilepsy patient with TI stimulation in Fig. 7. First, we used high resolution modeling to simulate TI stimulation. We calculated the TI electric field, which is projected onto the cortical surface by stimulating two pairs of scalp-EEG electrodes AFz-Fpz and AF3-Fp1 in Fig. 7**A**) at the frequencies of 1000 Hz and 1005 Hz, respectively. The brain source signals on each vertex of the whole brain is shown in (Fig. 7**B** and Movie S4). We then mapped the neural activity from the cortical surface onto the SEEG and scalp-EEG electrodes (Fig. 7**C**) using the source-to-sensor mapping matrix. We extracted the data features from simultaneous ictal synthetic SEEG and scalp-EEG recordings (Fig. 7**C**) which served as the input to the multimodal model inversion modules to estimate the EZN. We obtained the left middle frontal cortex as the region with highest EVs from all samples (Fig. 7**D**), shown in red in 3D in Fig. 7**E**). The left F1-lateral prefrontal and left-Orbito-frontal-sulcus were identified as the second candidates belonging to the EZN (Fig. 7**D**) and are shown in yellow in Fig. 7**E**.

**Fig. 7.**
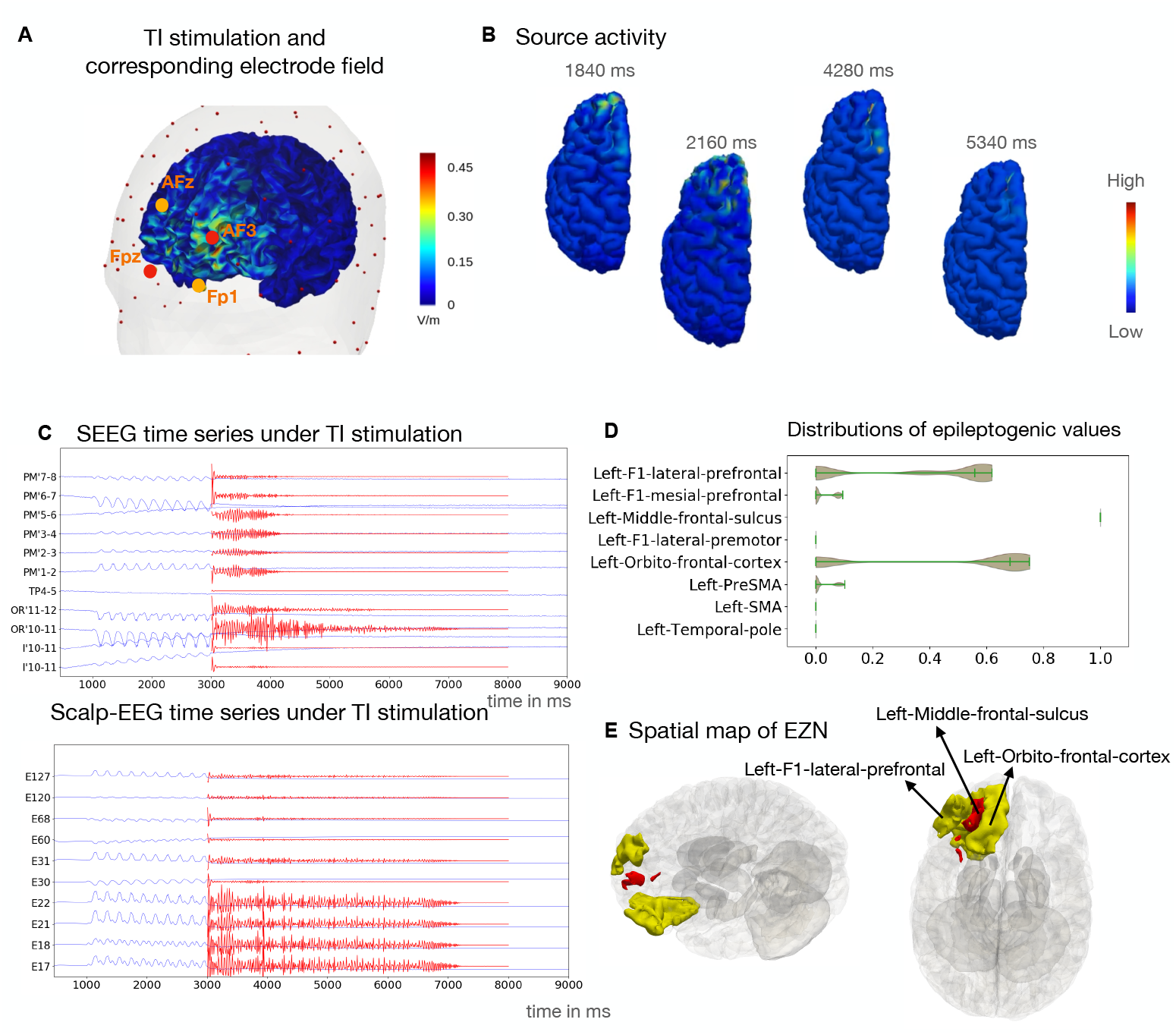
Estimating EZN from TI stimulation for a second patient with frontal lobe epilepsy. (**A**) The electric field of TI stimulation by two pairs of scalp-EEG electrodes (shown in red and orange). We applied a frequency of 1000 and 1005 Hz for the first and second electrode pairs, respectively. The Spatial distribution of the peak activity of the TI envelope is colored in the 3D brain. (**B**) Seizure dynamics were simulated using the Epileptor-Stimulation model through the TI stimulation. Neural activity is shown on the cortical mesh at 4 different time points. (**C**) Selected synthetic SEEG time-series (top) and scalp-EEG time-series (bottom) from TI stimulation-induced seizure. The scaled-up time series during the seizure period is shown in red. (**D**) Posterior distribution of the EV (higher value indicates higher chance for seizure) for 8 selected regions the same as Fig. 6 **D**. (E) The highest chance of being the EZN as left-middle-frontal-sulcus in red mapped in 3D brain and left-F1-lateral-prefrontal and left-orbito-frontal-cortex in yellow.

## 3 Discussion

### 3.1 Summary

We developed a virtual brain twins pipeline tailored for stimulation based on personalized whole brain network modeling in epilepsy. This pipeline integrates empirical patient data and undergoes validation using simulated data generated from high-resolution network modeling. The high-resolution virtual brain twin models are able to explore stimulation effects. Moreover, the proposed pipeline employs model inversion through HMC and Bayesian inference to estimates EZNs. We demonstrated the estimation of the EZN from SEEG and scalp-EEG signals during seizure activity triggered by direct electrical SEEG or TI stimulation. We introduced multimodal model inversion methods integrating multiple, potentially simultaneous, modalities as well. The virtual brain twins pipeline for stimulation provides a directly applicable methodological and conceptual basis for a series of scientific studies and clinical translations. The concept of virtual brain twins from this pipeline can be extended to other brain disorders, such as Alzheimer’s and psychiatric disorder, among others [17]. In the subsequent discussion, we will delve into its methodological and conceptual implications for practical applications.

### 3.2 Future scientific studies

The first application of this pipeline can help us answering a crucial diagnostic question: How to infer the underlying EZNs from stimulated seizures? To answer this question, it is necessary to investigate the following problems, illustrated in Fig. S4. First, what is the proper range of stimulation parameters to induce seizures for diagnosis? Currently, no standardized stimulation protocols exist [20]. The French guidelines suggest parameters for both low-frequency (1 Hz) and high-frequency (50 Hz) stimulation with specified ranges on pulse width, pulse intensity and stimulation duration [3]. Our personalized whole brain model has the ability to predict the stimulation effect for individual patients given specified stimulation parameters and should be validated in future empirical studies. On the patient-specific level, among all feasible stimulation parameters, then what are the optimal stimulation parameters for inducing seizures for diagnosis? Second, what is the relationship between the EZN triggered by stimulation and the one occurring during spontaneous seizures? More specifically, we want to investigate whether the EZN estimated from stimulation induced seizures equals the EZN of spontaneous seizures. It could be possible that stimulation forces seizures in brain regions that would not be epileptogenic otherwise, or that stimulation uncovers only a subset of epileptogenic regions. Even for spontaneous seizures, we have to remember that the limited observations of recordings of spontaneous seizures may also only tell part of the story. Third, how many induced seizures by SEEG stimulation are sufficient for identifying the full underlying EZN? We can investigate this question by systematically evaluating the EZNs on SEEG stimulated induced seizures generated by virtual brain twin models with given EZNs. The proposed VEP stimulation pipeline with high-resolution modeling provides a reasonable framework as a useful tool to address all these questions.

The proposed virtual epilepsy patient pipeline has the capability to integrate multimodal neuroimaging data, including anatomical recordings, such as T1-weighted MRI, diffusion-weighted MRI, and CT; as well as functional ones, such as scalp-EEG, MEG, SEEG, subdural grid, PET and fMRI. The framework that we introduced here takes this process further and the multimodal inference demonstrates possible approaches to integrate information from scalp-EEG and SEEG. This process can be easily extended to other recordings, such as MEG, fMRI, and subdural grid electrodes, depending on the available data. The only difference between the different functional measures is the forward solution matrix. We designed an multimodal inference version of the HMC algorithm, which can be directly used for simultaneous functional recordings, such as SEEG/scalp-EEG [30] or MEG/scalp-EEG [31] or SEEG/MEG [32]. The proposed pipeline also provides a basic structure for estimating the EZN using interictal spikes as a datafeature from interictal SEEG/MEG recordings. The usage of this multimodal inference is also valid in cases where different recordings start with the same brain states, or when the sampling algorithms can cover all possible initial conditions.

The diagnostic pipeline for TI stimulation can provide a theoretical basis for non-invasive diagnosis and treatment of epilepsy. TI stimulation is a good candidate for both diagnosis and treatment in epilepsy within the non-invasive stimulation family. Other non-invasive stimulation techniques, such as tDCS [33], are mostly used for treatment only in epilepsy. The key feature of TI stimulation is that it enables a focused stimulation which can reach deep brain structures non-invasively [24, 34], such as the hippocampus, which quite often is involved in the EZN in temporal lobe epilepsy. Studies have demonstrated the safety of TI in human participants [35]. Literature and seminars with preliminary results have started to discuss the application of TI stimulation to brain disorders, such as epilepsy [23, 36], Alzheimer’s [37], Parkinson’s [38] and essential tremor [39]. TI stimulation has evoked seizure-like events in the mouse hippocampus that have been experimentally well controlled [40]. Our study here provides a pipeline for predicting the use of TI for the diagnosis of the EZN in human epilepsy, and as a basis for an optimization strategy for TI stimulation parameters. Based on this proposed framework, we can further build high resolution models for individual patients in future therapeutic interventions. Although we obtained a good estimate of the EZN from non-invasive scalp-EEG signals for patient examples whose EZNs were in cortical regions, for future non-invasive diagnosis of EZNs in deep structures, we need to explore the non-invasive measurements that can provide useful data features from these deep brain structures.

The current pipeline has several limitations. Model-based approaches are usually computationally expensive and parameter sensitive, which may pose challenges for real-time use in clinical routine, such as the optimization of stimulation parameters. The current pipeline does not consider the dynamics of the epileptic disorders over long timescales nor the long-term effects of the stimulation. Ongoing systematic evaluations and studies will also improve the current pipeline in the following ways. High-resolution modeling of the subcortical structure and a more realistic forward resolution may improve the whole brain modeling. The introduction of regional variations (such as cell density and receptor density) [41, 42] may provide a fundamental way to improve the current pipeline.

## 4 Methods and Materials

### 4.1 Patient data

We used the data from two patients who underwent a standard presurgical protocol at La Timone Hospital in Marseille. Informed written consent was obtained in compliance with the ethical requirements of the Declaration of Helsinki and the study protocol was approved by the local Ethics Committee (Comité de Protection des Personnes sud Méditerranée 1). This patient underwent a comprehensive presurgical assessment, which included medical history, neuropsychological assessment, neurological examination, fluorodeoxyglucose positron emission tomography, high-resolution 3T MRI, long-term scalp-EEG and invasive SEEG recordings. The patient received non-invasive T1-weighted imaging (MPRAGE sequence, repetition time = 1.9 *s*, echo time = 2.19 *ms*, voxel size 1.0 × 1.0 × 1.0 *mm*^3^) and diffusion-weighted MRI images (with an angular gradient set of 64 directions, repetition time = 10.7 *s*, echo time = 95 *ms*, voxel size 1.95 × 1.95 × 2.0 *mm*^3^, b-weighting of 1000 *s/mm*^2^.) The images were acquired on a Siemens Magnetom Verio 3T MR-scanner. The patient had invasive SEEG recordings obtained by implanting multiple depth electrodes, each containing 10−18 contacts 2 *mm* long and separated by 1.5 or 5 *mm* gaps. The SEEG signals were acquired on a 128 channel Deltamed/Natus system. After the electrode implantation, a cranial CT scan was performed to obtain the location of the implanted electrodes.

### 4.2 Data processing

To construct the individual brain network models, we first preprocessed the T1-MRI and diffusion-weighted MRI data. Volumetric segmentation and cortical surface reconstruction were from the patient-specific T1-MRI data using the recon-all pipeline of the FreeSurfer software package http://surfer.nmr.mgh.harvard.edu. The cortical surface was parcellated according to the VEP atlas [26], the code for which is available at https://github.com/HuifangWang/VEP_atlas_shared.git. We utilized the MRtrix software package to process the diffusion-weighted MRI [43], employing the iterative algorithm described in [44] to estimate the response functions and subsequently used constrained spherical deconvolution [45] to derive the fiber orientation distribution functions. The iFOD2 algorithm [46] was used to sample 15 million tracts. The structural connectome was constructed by assigning and counting the streamlines to and from each VEP brain region. The diagonal entries of the connectome matrix were set to 0 to exclude self-connections within areas and the matrix was normalized so that the maximum value was equal to one. We obtained the location of the SEEG contacts from post-implantation CT scans using GARDEL (Graphical user interface for Automatic Registration and Depth Electrodes Localization), which is part of the EpiTools software package [47]. Then we coregistered the contact positions from the CT scan space to the T1-MRI scan space for this patient.

### 4.3 High resolution simulation of the Epileptor-Stimulation model

We modeled epileptic seizures on a patient-specific high-resolution brain model. Although there is no consensus about the precise biophysical mechanism of ion exchanges that leads to seizure onset. Previous studies demonstrated an increase in excitability, spike frequency and oscillation power [48–51] when an external perturbation was applied. Experimental studies have linked seizure onset to specific changes in ion dynamics such as extracellular potassium or intracellular chloride [52–54]. We hypothesized that exposure of repetitive stimulation/pertubation generates a slow accumulation effect, which can push the system to the seizure state when a seizure threshold is reached.

We model this accumulation effect phenomenologically via the parameter *m*, which influences the oscillatory dynamics in the seizure onset state in the Epileptor model [55, 56]. We render *m* time dependent, *m* = *m*(*t*), and introduce an exponentially decaying memory kernel of length *r* through a linear ordinary differential equation, which slowly builds up when an external stimulus is applied until the seizure threshold is reached. In the absence of a stimulus, *m*(*t*) slowly returns back to baseline. *H* is the Heaviside function. When *m* is greater than the seizure threshold *m*_*thresh*_, the *H* function equals 1, which kicks the system into the seizure-like state. Otherwise, the *H* function equals 0.

The pial surfaces of both hemispheres define the spatial domain along which the network activity can unfold. Neural dynamics are governed by an extension of the original Epileptor model [25]. The extended Epileptor-Stimulation model includes a stimulation-accumulating state variable *m* that can destabilize the system and produce a seizure. The neural activity at every vertex *i* = 1, …, *N* of the network is governed by the following equations:

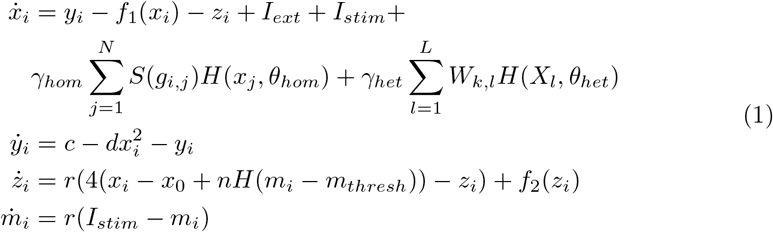

where:

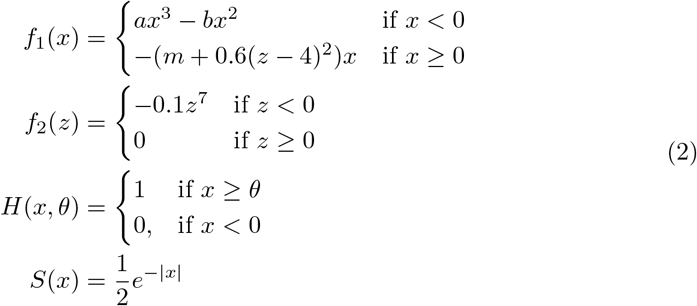

The state variables *x* and *y* describe the activity of neural populations on a fast time scale and can model fast discharges. The oscillation of the slow permittivity variable *z* drives the system autonomously between ictal and interictal states. The parameter *x*_0_ indicates the degree of excitability and directly controls the dynamics of the neural population to produce seizures or not. Each vertex *i* is connected locally through homogeneous connections and globally through heterogeneous connections to other parts of the brain. Although local connections are actually heterogeneous with a complex variability of interneurons and dendritic arborization, due to lack of personalized measurement in clinical routines, we made an assumption of homogeneity in the local connections while considering only geometric features of the cortical surface. Local connections, parameters with the subscript *hom*, are described by a translationally invariant Laplacian coupling kernel *S*(*g*_*i,j*_) where *g*_*i,j*_ denotes the geodesic distance along the cortical mesh between vertices *i* and *j*. Additionally, global connections, parameters with the subscript *het*, along white matter fibers, are accounted for by the connectome matrix, where *W*_*k,l*_ is the connection strength between brain areas *k* and *l*. Each vertex *i* is assigned to a brain area *k* = 1, …, *L* according to a cortical parcellation. For this study we used the VEP parcellation. The average neural activity *X*_*l*_ of all vertices belonging to area *l* is coupled throughout the network and projected uniformly to all vertices of area *k*. The default parameters of the system are *I*_*ext*_ = 3.1, *c* = 1, *d* = 5, *r* = 1, *a* = 1, *n* = 1, *m*_*thresh*_ = 1.8, *γ*_*het*_ = 0.1, *γ*_*hom*_ = 0.8, *θ*_*het*_ = −1, *θ*_*hom*_ = −1. The time varying input *I*_*stim*_ describes the perturbation signal in each time step depending on stimulation parameters. To accomplish this, we computed the generated electric field from the two types of perturbation a) an invasive SEEG stimulation, and b) a non-invasive TI stimulation. These are described in two separate subsections below.

### 4.4 Calculation of the electric field of SEEG stimulation

The SEEG stimulation is applied to a pair of neighboring sensors, in which one acts as a cathode and the other one as an anode. This generates a bipolar pulse perturbation in the area where the electrodes are located. However, the effects of the stimuli are not only local, because they can propagate through short-range and long-range connections towards other brain areas. The parameters used clinically are restricted to frequencies of either 1 Hz or 50 Hz, weak amplitudes ranging from 0.1 to 3 mA, and pulse widths of 500 – 2000 microseconds. We modeled the same stimulus as had been applied clinically on a personalized digital brain model of the patient. Using the reconstructed brain structure and the overlayed SEEG electrodes, we generated a biphasic signal using the stimulation parameters for the pair of electrodes chosen by the clinician. We then mapped the stimulus signal onto the parcellated brain areas based on the distance, which resulted in an estimated electric field across the brain regions. We used this estimated field as an input to the *I*_*stim*_ parameter and simulated the time series on selected vertices from the EZ.

### 4.5 Calculation of the electric field of the TI stimulation

Non-invasive stimulation with TI is a recent method that aims to mimic DBS’s capabilities to be both focal and sub-cortical without being invasive. This is done using the assumption that high frequencies (above 500 Hz) have an ignorable effect on neuronal activity [24, 34]. By combining multiple fields of very high-frequency tACS stimulations, such that they interfere maximally in the target of choice, a new electric field can be generated, which modulates at much lower frequencies (e.g. two fields of 1000 and 1050 Hz modulate at 50 Hz at the target of choice). The generated electric field was computed using SimNIBS, as described previously, to compute the generated tACS fields. Then, the vectorial tACS fields are added together, resulting in a TI field estimate for the brain regions. We used this estimated field as an input to the *I*_*stim*_ parameter and simulated the time series on one selected vertex from the EZ.

To obtain the distribution of the TI field in the gray matter, we started by computing the electric field distribution from the simulation of a transcranial direct current stimulation (tDCS) via SimNIBS [57]. First, we reprocessed the individual T1-weighted MRI via the SimNIBS process “mri2mesh”, which resulted in the segmentation into five head tissues: white and grey matters, cerebrospinal fluid, skull and scalp. This process creates the tetrahedral meshes necessary for the finite element method (FEM) for the simulation of the electric field distribution. We computed the tDCS field for each pair of electrodes positioned based on the location of the coregistered 128 electrodes scalp-EEG cap, using the 10-5 system. Circular electrodes with a diameter of 10 mm and a thickness of 5 mm were directly applied on the head. Then, we projected these electric fields onto the resampled Freesurfer surface (20484 vertices). We extracted the amplitude of the electric field at each vertex of the cortex surface. These fields derived from the coupled electrodes pairs were then modulated by multiplying them with two sinusoids with the frequency of 1000 Hz and 1005 Hz, respectively, for 1 second. Two oscillatory electric fields, which were linearly summed, provided an interference field. To obtain a correct representation of the high frequency activity, the sampling rate was 30 kHz. We extracted the 5 Hz peak envelope by using spline interpolation. Finally, we included this envelope in the virtual brain model as the source of external simulation for the cortex vertices (20484). For this patient, we targeted the highest TI stimulation effect on the left O2 in the VEP atlas [26]. We used the optimization process in SimNIBS to identify the best 4 electrodes configurations (10-5 scalp-EEG montage), which maximized the electric field in the left O2. This procedure allowed us to identify, in our case, PPO3-PPO5 and P5h-PO5h ([28]) as the best electrodes couple for the left-O2 stimulation.

### 4.6 Forward solution for SEEG signals

The forward solution for the SEEG signals maps the neural activity from the sources to the sensors (SEEG contacts), represented by a source-to-sensor matrix (gain matrix). As sources for our model, we used the vertices of the pial surface for the cortical regions, and each subcortial region as a single node as in the neural mass model (NMM). Surfaces are represented as triangular meshes. For the NMM, we defined the mapping *g*_*j,k*_ from the source brain region *j* to the sensor *k* as the sum of the inverse of the squared Euclidean distance *d*_*i,k*_ from vertex *i* to sensor *k*, weighted by the area *a*_*i*_ of the vertex on the surface.

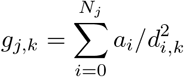

Here vertex *i* belongs to region *j* which has *N*_*j*_ vertices in total. The area *a*_*i*_ of vertex *i* was obtained by summing up one third of the area of all the neighboring triangles. Vertices belonging to the same brain region were summed to obtain the gain for a single region of our brain network model. The resulting gain matrix has dimensions *M* × *N*, with *M* being the number of regions and *N* the number of sensors.

Matrix multiplication of the simulated source activity with the gain matrix yielded the simulated SEEG signals, i.e., *SEEG*_*k*_(*t*) = ∑_*j*_ *g*_*j,k*_*x*_*j*_ (*t*), where *x*_*j*_ (*t*) is the time series of the source level signals. This distance-based approach and the summation of all the vertices within each region neglect the orientation of the underlying current dipoles. Pyramidal neurons, which are oriented normal to the cortical surface, are assumed to be the physiological source of any electric signal recorded with SEEG, scalp-EEG, or MEG [58]. The direction of the NMM is ill defined a t typical spatial resolutions of 10-20 *cm*^2^ in virtual brain networks of 100-200 nodes [26]. However, approaches that use high resolution representation of the network, allowing to compute an surface orthogonal, have this ability. To solve the forward problem we followed the analytical solution proposed in [59] for electric fields i n a n u nbounded homogeneous medium. This choice of forward model assumes no boundary effects of changes of conductivity at tissue boundaries. A previous study has shown that the error of an unbounded homogeneous conductivity model compared to a more accurate finite element method model with changes in conductivity is relatively small for electric fields generated by dipoles deep in the brain and electrodes close to the source [60]. Therefore, we can estimate the gain matrix elements *g*_*i,k*_ for the NMM approach by

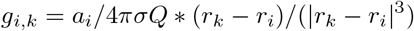

where *r*_*k*_ and *r*_*i*_ are the position vectors of sensor *k* and source vertex *i*, respectively. |*v*| represents the L2 norm of a vector *v. Q* is the dipole orientation vector, and *σ* is the electric conductivity. Since we assume constant conductivity across the brain, it becomes merely a scaling factor, which we set to *σ* = 1.

### 4.7 Forward solution for scalp-EEG signals

To compute the forward solution of scalp-EEG signals, we first reconstructed three individual surfaces (inner skull, outer skull, and head) of the patient based on boundary element models (BEM) using Brainstorm [61]. Then in Brainstorm, we coregistered the scalp-EEG electrodes positions (of the Hydrocel E1 128 channels electrode cap) onto the head surface, according to the fiducial points of the patient’s T1-MRI. We applied a slight manual correction to better orient the scalp-EEG cap to the individual anatomy. Finally, we derived an scalp-EEG forward model using a 3-shell BEM model (conductivity: 0.33, 0.165, 0.33 S/m; ratio: 1/20) [62] and the OpenMEEG method implemented in Brainstorm [63, 64], to provide a realistic head model. Then we obtained a gain value for each dipole (20484 vertices), with the constrained direction normal to the cortex surface, for each scalp-EEG electrode. Finally, the gain matrix derived from the head model was multiplied by the simulated time series of the brain. sources to obtain the scalp-EEG activity, i.e. 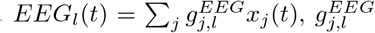 from the source signal on brain region *j* to the *EEG* signal on channel *l*

### 4.8 Calculation of the SEEG/scalp-EEG data features

We extracted the data features from the SEEG and scalp-EEG signal to be the input of the model inversion modules. The SEEG data was re-referenced using a bipolar montage, which was obtained using the difference between 2 neighboring contacts on one electrode. The 2D Epileptor model, introduced below, is suitable for fitting the envelope of the seizure time series. Ideally, the envelope follows a slightly smoothed rectangular function from the onset until the offset of the seizure. To get a well-formed target that our model should fit, we extracted the bipolarized SEEG signal from 10s before the seizure onset until 10s after the seizure offset. We identified the outlier time points that were greater than 2-times the standard deviation of the extracted signal and replaced them with the mean of the extracted signal. The signal was then high-pass filtered with a cut-off at 10 Hz to remove slow signal drifts. The envelope was calculated using a sliding-window approach with a window length of 100 time points. The signal inside the window was squared, averaged and log transformed. From the resulting envelope, we again identified and removed outliers, as described above. Finally, the envelope was smoothed using a lowpass filter with a cut-off of 0.05 Hz. The mean across the first few seconds of the envelope was used to calculate a baseline, which was then subtracted from the envelope. The same procedure was used for the EEG data, the only difference being that we used absolute values for the gain matrix, in order to get the accumulated effect on each EEG electrode throughout the entire seizure.

### 4.9 The HMC model inversion

By taking advantage of time scale separation and using averaging methods, the Epileptor can be reduced to a 2D system [65]:

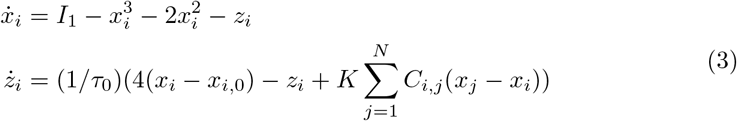

where *τ*_0_ scales the length of the seizure. The external input is defined as *I*_1_ = 3.1. We used the 2D Epileptor for the model inversion, i.e. the parameter estimate of the model, from the scalp-EEG and SEEG recordings.

For the individual SEEG model inversion, the forward solution is: *SEEG*_*k*_(*t*) = ∑_*j*_ *g*_*j,k*_*x*_*j*_(*t*). For the individual scalp-EEG model inversion, the forward solution is: 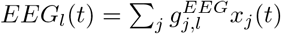.

For the multimodal model inversion, we projected the same source signals to the scalp-EEG and SEEG at the same time while we calculated the likelihood according to the HMC algorithm, i.e. *SEEG*_*k*_(*t*) = ∑_*j*_ *g*_*j,k*_*x*_*j*_(*t*); 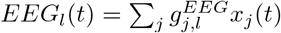.

For the model inversion, we applied the No-U-Turn Sampler (NUTS), an adaptive variant of the HMC algorithm to sample the posterior density of the model parameters. The performance of the HMC is highly sensitive to the step size and number of steps in the leapfrog integrator for updating the position and momentum variables in a Hamiltonian dynamic simulation [66]. We used NUTS, which is implemented in Stan and extends HMC with adaptive tuning of both the step size and the number of steps in a leapfrog integration to sample efficiently from the posterior distributions [66, 67]. To overcome the inefficiency in the exploration of the posterior distribution of the model parameters, we used a reparameterization of the model parameters based on the map function from the model configuration space to the observed measurements [10]. Our reparameterization-based approach reduces the computation time by providing more effective sample sizes and removing divergences by exploring the posterior distributions of the linear combinations of regional parameters that represent the eigenvectors obtained from the singular value decomposition of the gain matrix. We denote the matrix of the eigenvectors of *G*^*T*^ *G* as *V* and the new parameters as 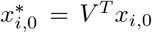 and *z*_*i*_(*t*_0_)^*^ = *V* ^*T*^ *z*_*i*_(*t*_0_). We ran the model inversion on both empirical and simulated seizures with 16 chains starting from eight optimized initial conditions. The eight optimized initial conditions are the output of maximum a posterior estimation algorithm [15]. We ran the maximum a posterior estimation algorithm 50 times and selected the best eight results in terms of the likelihood. We assessed model identifiability based on an analysis of posterior samples, which demonstrated that the sampler explores all the modes in the parameter space efficiently. The analysis includes trace-plots (evolution of parameter estimates from draws over the iterations), pair plots (to identify collinearity between variables), and autocorrelation plots (to measure the degree of correlation between draws of samples). Sampling convergence of the algorithms was assessed by estimating the potential scale reduction factor and calculating the effective sample size based on the samples of the posterior distributions, providing estimates of the efficient run times of the algorithm.

### 4.10 Data, Materials and Software Availability

. All shareable codes and data are included in the manuscript and/or Supplementary documents.

## 5 Fundings

The preparation of this article was funded through the EU’s Horizon Europe Programme under grant No. 101147319 (EBRAINS 2.0) and No. 101137289 (Virtual Brain Twin), The Amidex Recherche Blanc under grant, No. AMX-22-RE-AB-135 (HR-VEP).

## Supporting information

All supplemental figures

## Data Availability

All data produced in the present study are available upon reasonable request to the authors.

## Notes

### Competing Interest Statement

The authors have declared no competing interest.

### Author Declarations

We used the data from two patients who underwent a standard presurgical protocol at La Timone Hospital in Marseille. Informed written consent was obtained in compliance with the ethical requirements of the Declaration of Helsinki and the study protocol was approved by the local Ethics Committee (Comité de Protection des Personnés sud Mediterranée 1).

