## Supplementary material for "Virtual brain twins for stimulation in epilepsy": All supplemental figures: VEP_stimulation_sup.pdf

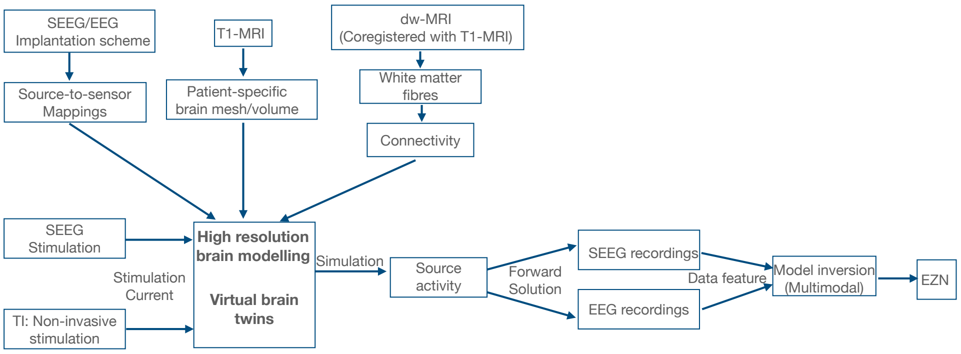


**Fig. A1 Flowchart of the virtual-brain-twin stimulation pipeline**. First, a T1-MRI defines the patient-specific high-resolution space. Second, the structural connectivity, the gain matrix (source-to-sensor matrix), and sensor-to-source matrix are obtained in a patient-specific brain space by co-registering both diffusion-weighted MRI and CT for SEEG locations and standard EEG system configuration with T1-MRI. We extract data features from SEEG recordings and EEG recordings from both SEEG-stimulation and TI stimulation and use HMC techniques for model inversion. From each stimulation module we can obtain a posterior distribution of epileptogenic values, which suggests EZN candidates. Then the integration module combines multiple results in different cases to help the clinical decision.

### A.1 movies

Movie 1: The brain activity of Patient 1 under SEEG stimulation on high-resolution brain surfaces. The bipolar SEEG stimulation initiates at 505 ms, lasting for 3 seconds. A seizure is induced around 3,505 ms and terminates around at 10,255 ms. Observing seizure activity at the source level can help our understanding epileptogenic zones.

Movie 2: The brain activity of Patient 1 under temporal interference stimulation on high-resolution brain surfaces. The temporal interference stimulation initiates at 500 ms, lasting for 6 seconds. A seizure is induced at approximately 3,505 ms and terminates around 13,400 ms.. Observing seizure activity at the source level can help our understanding epileptogenic zones.

Movie 3: The brain activity of Patient 2 under SEEG stimulation on high-resolution brain surfaces. The bipolar SEEG stimulation initiates at 504 ms, lasting for 3 seconds. A seizure is induced around 3,504 ms and concludes at 10,072 ms. Observing seizure activity at the source level can help our understanding epileptogenic zones.

Movie 4: The brain activity of Patient 2 under temporal interference stimulation on high-resolution brain surfaces. The temporal interference stimulation initiates at 1,000 ms, lasting for 2 seconds. A seizure is induced around 3,000 ms and concludes at 7,660 ms. Observing seizure activity at the source level can help our understanding epileptogenic zones.

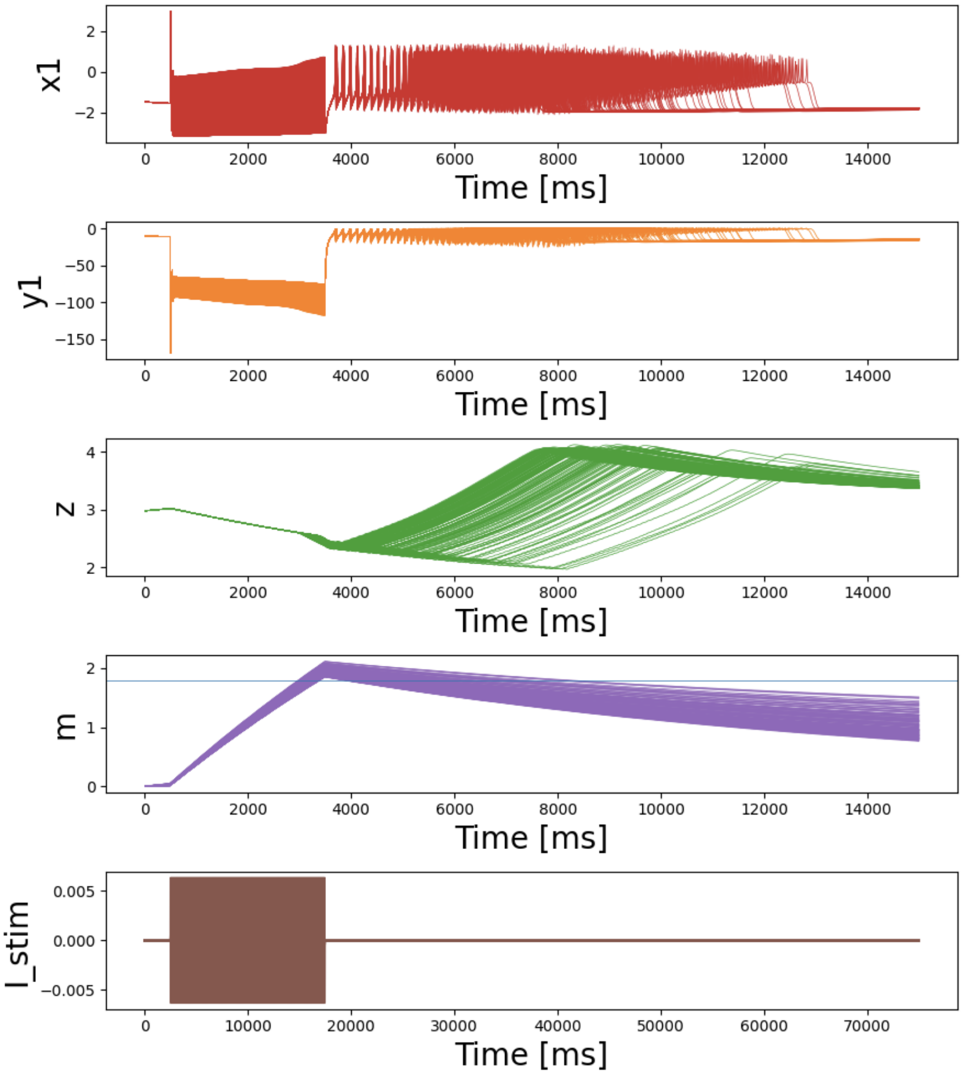


**Fig. A2** The time-series of Epileptor stimulation variables during induced seizures through SEEG stimulation. Five state variables of the model at seizing parts of the cortex and the stimulation time course $I_{stim}$. The stimulation is accumulating on state variable $m$ of the Epileptor-Stimulation equation 1. Once $m$ passes a fixed threshold it causes the model to initiate the seizure.

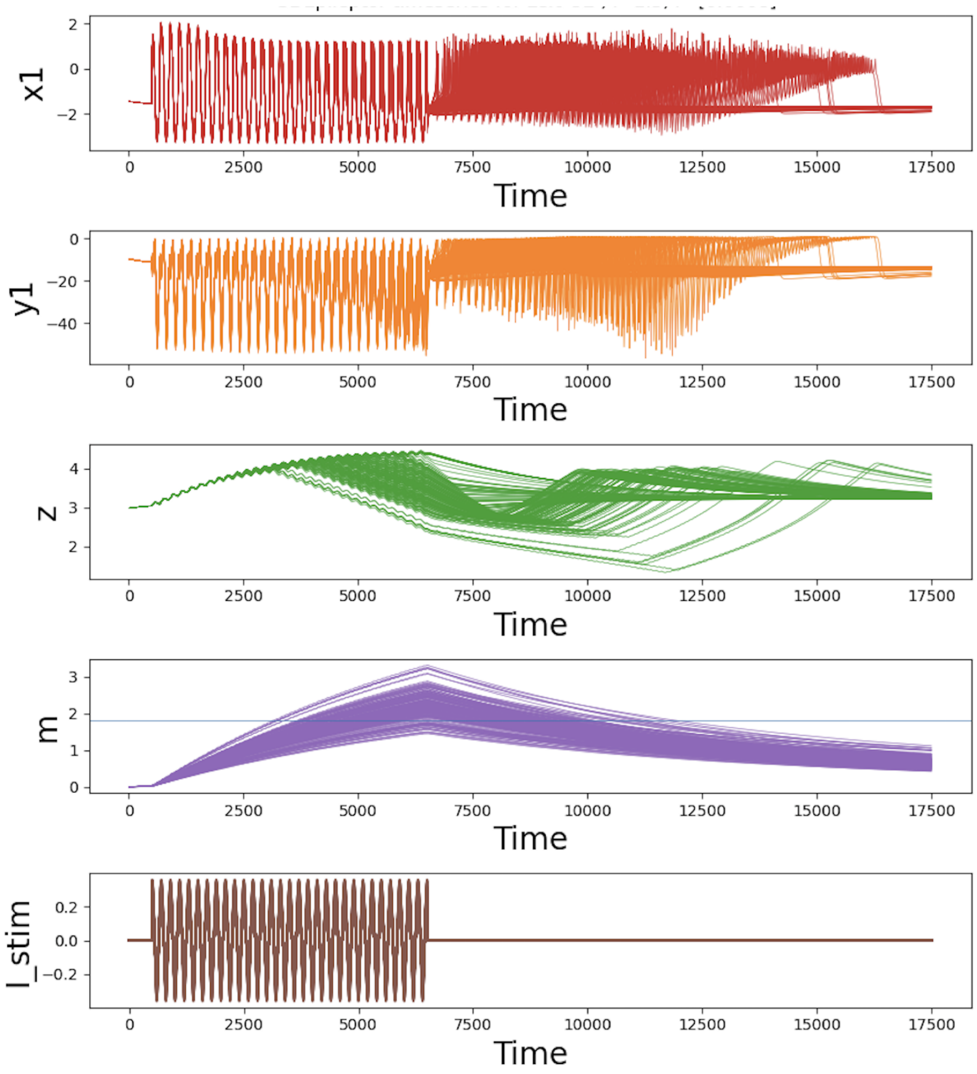


**Fig. A3** The time-series of Epileptor-Stimulation equations by inducing seizure through stimulation induced by TI. Five state variables of the model at seizing parts of the cortex and the stimulation time course $I_{stim}$. The stimulation is accumulating on state variable m of the Epileptor. Once m passes a fixed threshold it causes the model to initiate the seizure.

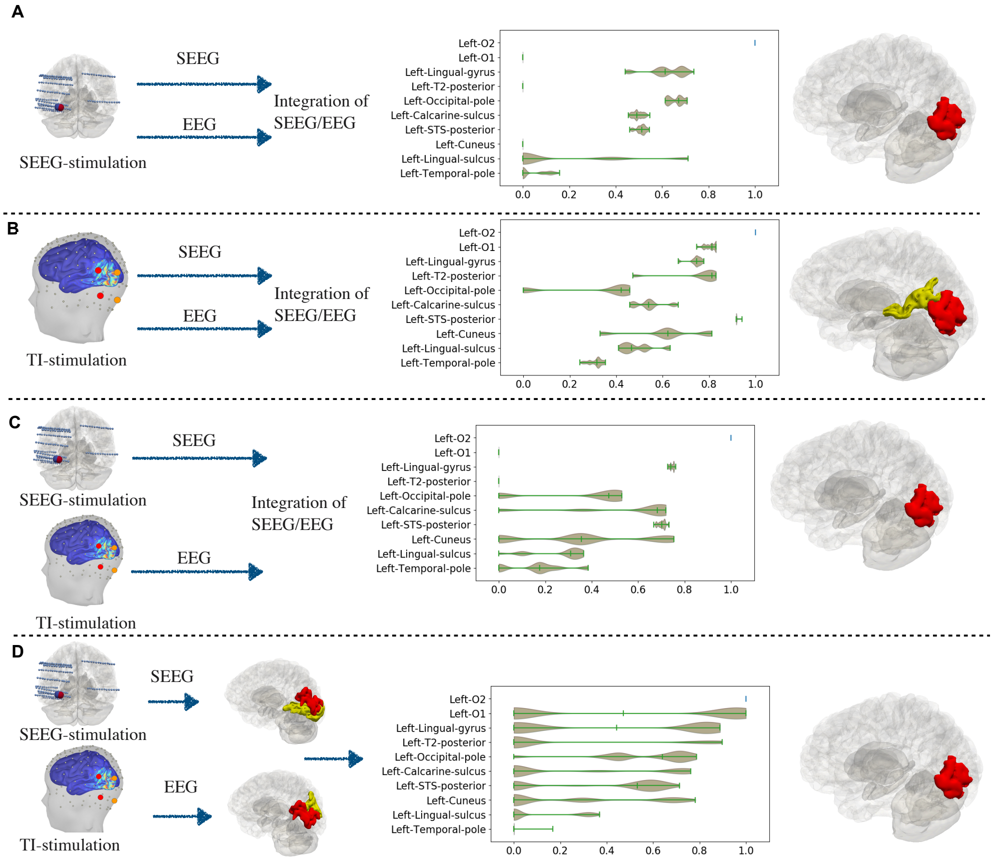


**Fig. A4** Integration module for multiple SEEG and scalp-EEG recordings.(**A-C**) The posterior distribution of EVs from using the HMC on a simultaneous SEEG and scalp-EEG data, which were mapped from the same high resolution source data. This model inversion can be performed in three cases: seizures induced by SEEG stimulation (**A**, the same case in figure 5**A**), by TI (**B**, the same case in figure 5**B**) and by both SEEG and TI (**C**). (**D**) A combination of the posterior distribution of EVs by pooling the EVs distribution obtained from SEEG (Figure 3D under SEEG stimulation) and scalp-EEG recordings (Figure 4D under TI stimulation). Each row shows the data used for model inversion, the posterior of the EVs and the corresponding highlighted EZs on the 3D brain, where the left O2 is in red and left STS-posterior is in yellow.

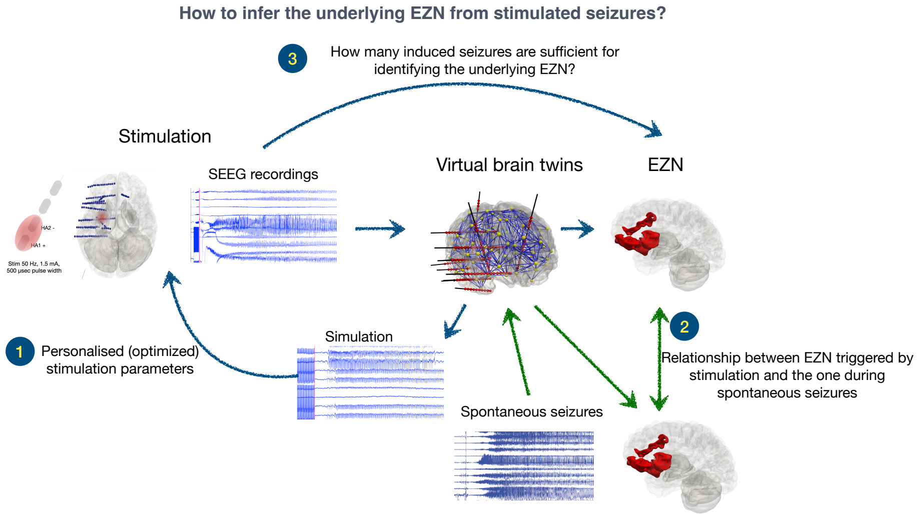


**Fig. A5** The study scheme to answer: How to infer the underlying EZNs from stimulated seizures? We illustrated three related systematic studies and details described in the first paragraph of subsection of future scientific studies.
